# Multidisciplinary stepwise treatments for lumbar disc herniation: a retrospective study

**DOI:** 10.1101/2024.12.10.24318769

**Authors:** Shaoting Zeng, Yan Weng, Ling Ye

**Affiliations:** Department of Pain Management, West China Hospital, Sichuan University, Chengdu, Sichuan Province, 610041, P. R. China; Department of Anesthesiology, The Peoples’ Hospital of Jianyang city, Sichuan Province, 641400, P. R. China; Department of Pain Management, West China Tianfu Hospital, Sichuan University, Chengdu, Sichuan Province, 610041, P. R. China

**Author notes:** Corresponding author: Department of Pain Management, West China Hospital, Sichuan University, Chengdu, Sichuan Province, 610041, P. R. China, Department of Pain Management, West China Tianfu Hospital, Sichuan University, Chengdu, Sichuan Province, 610041, P. R. China, Ling Ye. Shaoting and Yan Weng contributed equally to the manuscript. Source of support: Funded by grant 2023HXFH036 from 1·3·5 project for disciplines of excellence–Clinical Research Fund, West China Hospital, Grant 2023092 from Health Commission of Chengdu.

**Keywords:** Lumbar disc herniation, Pain management department, Orthopedics department, Rehabilitation department, Oswestry Disability Index, Multi-Disciplinary Treatment

## Abstract

**Objective:** The study was aimed to compare the efficacy of the treatment for lumbar disc herniation (LDH) in the pain management department, orthopedics department and rehabilitation department, and to explore the multidisciplinary stepwise treatments style.

**Methods:** This single-center retrospective study analyzed the clinical data from 1397 patients with LDH between June 2015 and July 2019 in the hospital. The patients were divided into three groups: Pain Management Department (P), Orthopedics Department (O), and Rehabilitation Department (R). Propensity score matching (PSM) was used to adjust for imbalanced confounding variables among the three groups. Patients’ general information, different style of treatments, visual analogue scale (VAS), duration of hospitalization, and hospitalization costs were recorded. Follow-up information of patients was obtained through the telephone, including: Oswestry dysfunction index (ODI), remission rate at discharge, the rate of three months revisit after the discharge. The independent student’s t test and chi-square test were applied to compare the differences among groups.

**Results:** After PSM, 144 patients from each group were included in the study and all covariates were well balanced among the three groups. In the matched patients, the order of remission rate at discharge was O>P>R(P<0.05), the rate of three months revisit after discharge was R (17.36%)> P (6.94%)>R (4.86%) (P <0.05). There was no significant difference in ODI index at discharge and follow-up between group O and group P(P> 0.05), while group R was higher than the other two groups (P <0.05). Patients in Group R had a longer length of hospital stay (P <0.05), while the hospitalization costs were ranked as O>P>R (P <0.05).

**Conclusions:** In the treatment of LDH, orthopedics department, pain management department and rehabilitation department could all achieve the relief of clinical symptoms, and the long-term efficacy was not stable. Patients presenting to the orthopedic department had the highest pain relief rate at discharge, low rate of the revisit at three months after discharge, followed by the pain management department and third by the rehabilitation department. We proposed that the treatment of LDH should be based on stepwise treatment and multidisciplinary treatment (MDT) model.

## Introduction

Lumbar disc herniation (LDH) is the main contributor to low back pain and sciatica, with a total incidence of 15-30% in Western countries and 18% in China[1]. LDH can cause chronic pain, especially the radiation pain of the lower limbs due to compression of the nerve root by herniated disc[2]. Recently, physic therapeutics, traditional Chinese medicine therapy, minimally invasive interventional surgery and surgical operation are the main treatments for LDH[3–6].Various previous studies and reviews have shown that the treatment of LDH follows a stepwise scheme of conservative treatment, minimally invasive treatment, and surgical treatment[7].In China, this stepped treatment involves the three most frequently treated departments for LDH patients: rehabilitation department, pain management department, and orthopedics department.

For the treatment of LDH, rehabilitation department adopts comprehensive treatment methods, including health education, exercise therapy, technique traction, acupuncture and drugs, etc. By reducing the pressure of the lumbar disc, enhancing the core muscle strength of patients, improving the local blood circulation, relieving muscle spasm, reducing the pain of patients to achieve the purpose of promoting patients’ rehabilitation. The pain management department primarily focuses on minimally invasive interventional therapy, utilizing physical or chemical methods act on the prolapsed intervertebral disc and compressed nerve roots, to reduce or remove the prominent disc, eliminate the edema of the compressed nerve to achieve the purpose of treating and relieving pain. Various treatments in pain management typically employ imaging devices such as X-rays and CT scans to guide procedures, ensuring the accuracy of drug injections while maximizing puncture safety and minimizing complications. With advantages such as smaller trauma and rapid postoperative recovery, minimally invasive interventional therapies have gained widespread clinical application[8–10]. Orthopedics department often chooses operations, including traditional open surgery and various minimally invasive endoscopic surgery to excise the diseased disc, removing its compression on the nerve root to achieve the purpose of treatment[11].

Recently, the above three departments make continuous progress in the treatment of LDH, and there are several studies and reviews on the diagnosis and specific treatment strategies in intervertebral disc diseases. However, previous studies showed that various treatment methods of LDH have recrudescence in in late-stage treatment, and most of the relevant researches reported the clinical efficacy of a single department in the treatment of LDH[12–14]. In this study, we expect that through the comparative analysis of the treatment effect of LDH in the three departments to provide a better diagnosis and treatment path of LDH, improving the medical quality and treatment effect.

## Materials and Methods

This was a single-center, retrospective study. We conducted a retrospective database review to patients whose first diagnosis were like "Lumbar Disc Herniation" at West China hospital from June 2015 to July 2019. This study was approved by the Ethics Committee of the West China hospital (No. 2019 – 965). This study is a retrospective data study, and the ethics committee waived the informed consent form. All patients were exempted from the signing of the informed consent form. The folw diagram of the study was shown in the figure 1.

**Figure 1.**
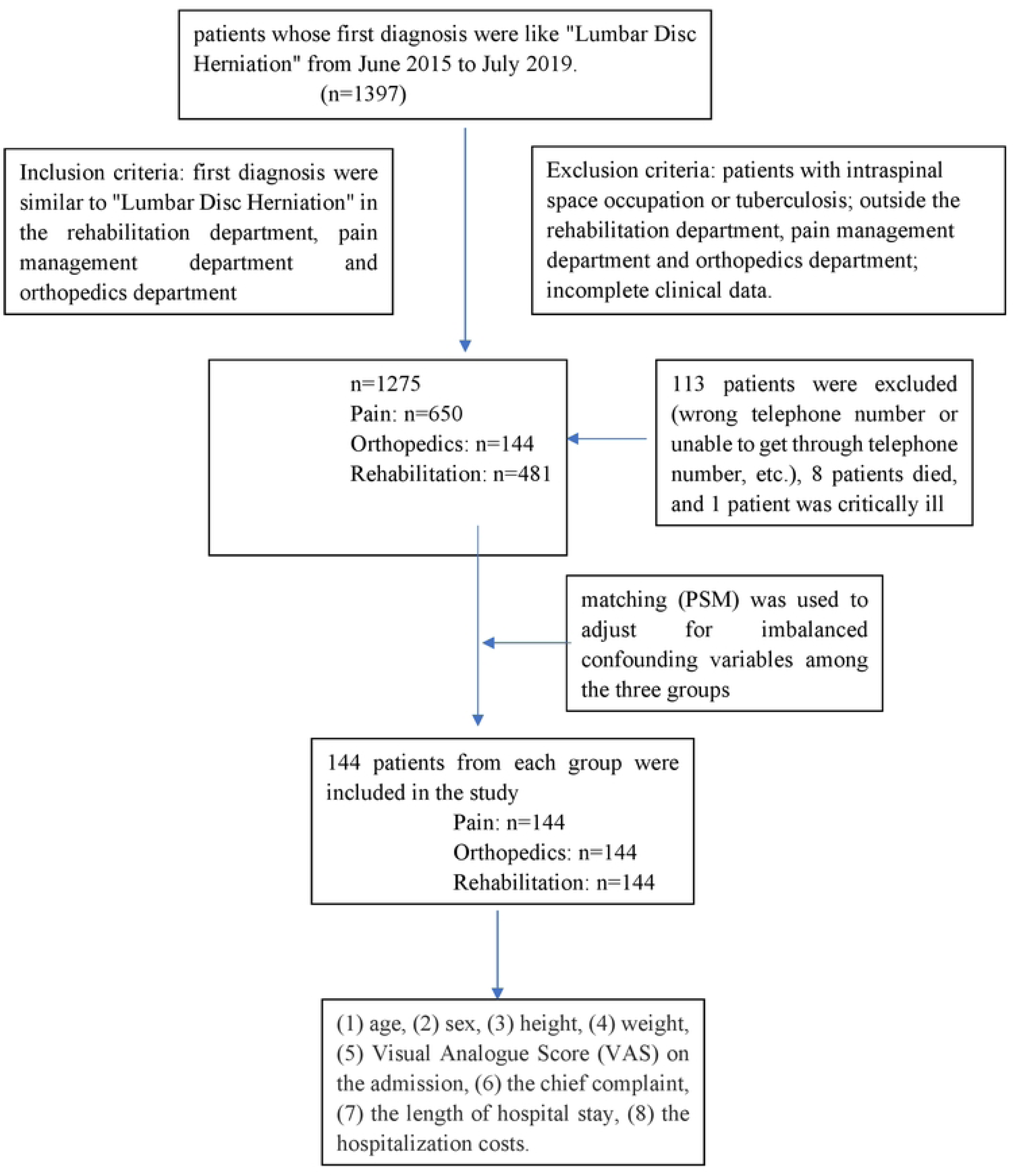
Flow diagram of the study

### Inclusion and Exclusion Criteria

Inclusion criteria were as follows: (i) patients whose first diagnosis were similar to "Lumbar Disc Herniation", diagnostic criteria were as follows: A)symptoms of low back pain and leg pain, lower extremity pain is typical of the lumbosacral nerve distribution area of pain and often more severe than low back pain; B)shows two of the four signs of nerve disorder: muscle atrophy, muscle weakness, abnormal sensation and reflex change; C) the straight-leg-raising test or femoral nerve stretching test was positive; D)imaging examination: abnormal signs of X-ray, CT, MRI are consistent with clinical manifestations;(ii) patients presenting to the rehabilitation department, pain management department and orthopedics department.

Exclusion criteria were as follows: (i) patients with intraspinal space occupation or tuberculosis; (ii) patients presenting outside the rehabilitation department, pain management department and orthopedics department; (iii) patients with incomplete clinical data.

After exclusion and inclusion criteria were fulfilled, there were 1397 patients with complete clinical data enrolled.

### Data Collection

The following pre-existing conditions already existed in the data base: (1) age, (2) sex, (3) height, (4) weight, (5) Visual Analogue Score (VAS) on the admission, (6) the chief complaint, (7) the length of hospital stay, (8) the hospitalization costs.

Follow-up information was obtained through the telephone, including the Oswestry Disability Index (ODI) at admission, the remission rate at discharge, the ODI at discharge, the ODI at follow-up, and the revisit at three months after discharge. Finally, the information of enrolled patients was exported through the database system for the statistical analysis.

Description of the data collection: This was a retrospective study, and the patients were not followed-up at a fixed time point. All enrolled patients were followed-up one time. The ODI was recorded using a telephone follow-up questionnaire. The remission rate at discharge was expressed as a percentage (%) by asking patients about their perceived remission of disease at discharge. Patients were asked whether they had returned to the hospital for LDH within three months after discharge.

### Observational indicators

Primary indicators: the ODI at admission, the ODI at discharge, the ODI at follow-up, the remission rate at discharge and the revisit at three months after discharge.

Secondary indicators: VAS on the admission; the length of hospital stay; the hospitalization costs.

### Statistical Analysis

Continuous variables were expressed as mean ± standard deviation (SD), T test or analysis of variance was applied to compare the differences if the data met the

parametric test conditions, otherwise, non-parametric tests were used. Categorical data were reported as count (constituent ratios) and comparisons between groups were analyzed by chi-square test. All statistical analyses were conducted in SPSS 23.0 and R 3.6.3 platform. P-values <0.05 were considered statistically significant.

## Results

### Patient Characteristics

A total of 1397 patients were included,113 patients were excluded (wrong telephone number or unable to get through telephone number, etc.), 8 patients died, and 1 patient was critically ill. Finally, 1275 patients were included in the analysis, including 650 patients from the Pain Management Department (P group), 144 patients from the Orthopedics Department (O group), and 481 patients from the Rehabilitation Department (R group). After initial exploratory analysis, there were significant differences in the baseline (age, VAS score, chief course of disease, ODI on admission) among P, O, and R groups. The baseline clinical characteristics of the three groups are detailed in Table 1(Table 1).

**Table 1.** Baseline characteristics of patients in the three groups.

| Characteristics | P(n=650) | O(n=144) | R(n=481) | P value |
| --- | --- | --- | --- | --- |
| Median age, years (range) | 62.00 (50.00-72.00) | 54.00 (45.00-65.00) | 52.00 (44.00-63.00) | 0.000* |
| Median BMI, Kg/m<sup>2</sup> (range) | 23.81 (22.04-25.95) | 24.22 (22.44-25.97) | 24.14 (22.05-25.65) | 0.322 |
| Median VAS scores, years (range) | 6.00 (5.00-8.00) | 2.00 (0.00-3.00) | 3.00 (2.00-4.00) | 0.000* |
| Median chief course of disease, months (range) | 24.00 (6.00-96.00) | 24.00 (6.00-120.00) | 12.00 (3.00-60.00) | 0.000* |
| Median ODI<br>on admission,<br>scores (range) | 18.00 (12.00-23.00) | 19.00<br>(15.00-25.00) | 15.00 (10.00-20.00) | 0.000* |

\* $p < 0.05$ means the corresponding confounders are not balanced among these groups

In order to unify the baseline of patients and minimize confounding bias in the three groups, we implemented a 1:1 PSM study. Currently, there is no internationally recognized indicator as the baseline uniform standard for LDH. ODI is the international common questionnaire index of back pain[15], this index can reflect the impact of lumbar pain on patients’ daily life. VAS score is a subjective index of patients in the international common pain degree evaluation. We did not include VAS score as a factor for propensity score matching because the three departments pay different attention to this index during the medical history collection, part of the data is missing. After PSM, 144 pairs were matched, and there were no statistical differences in age, sexual, Body Mass Index (BMI) and ODI at admission among the three groups (P> 0.05). The baseline characteristics of the three groups after PSM are detailed in Table 2 (Table 2).

**Table 2.** Baseline characteristics of the three groups after PSM.

| Characteristics | P(n=650) | O(n=144) | R(n=481) | P value* |
| --- | --- | --- | --- | --- |
| Median age,<br>years (range) | 53.00 (41.00-65.00) | 54.00 (45.00-65.00) | 52.50 (45.00-65.50) | 0.832 |
| Median<br>BMI,<br>Kg/m<sup>2</sup> (range) | 24.23 (22.06-26.31) | 24.22<br>(22.44-25.97) | 24.42 (22.68-26.21) | 0.988 |
| Median ODI<br>on admission,<br>scores (range) | 19.50 (12.00-25.00) | 19.00<br>(15.00-25.00) | 19.00 (14.00-25.00) | 0.451 |
| Female, sex, n<br>(%) | 67<br>(46.5) | 66<br>(45.8) | 59<br>(41.0) | 0.586 |

\* $p < 0.05$ means a significant difference among these groups.

### Discharge remission rate

There was statistical significance in the remission rate at discharge among the three groups (Table 3, P< 0.05). Pairwise comparison showed statistically significant difference in the remission rate of patients discharged from the three departments (Table 4, P< 0.05). The order of remission rate at discharge was O>P>R.

**Table 3.** remission rates at discharge in the three departments.

| Characteristics | groups | Mean value (95%CI) | P value |
| --- | --- | --- | --- |
| Discharge | P | 60.28 (57.18-63.38) | 0.000* |
| remission | O | 65.28 (62.44-68.12) |  |
| rate ,%(range) | R | 44.03 (40.80-47.25) |  |

\* $p < 0.05$ means a significant difference among these groups.

**Table 4.** Pairwise comparison of remission rates at discharge in the three departments.

| Characteristics | groups | groups | P value |
| --- | --- | --- | --- |
| Discharge | O | P | 0.000* |
| remission rate | O | R | 0.000* |
| (%) | P | R | 0.044* |

\* $p < 0.05$ means a significant difference among these groups.

### The rate of revisit after the discharge of three months

The rate of three months revisit after discharge was statistically significant (Table 5, P< 0.05), ranking as R (17.36%)> P (6.94%)>R (4.86%).

**Table 5.**
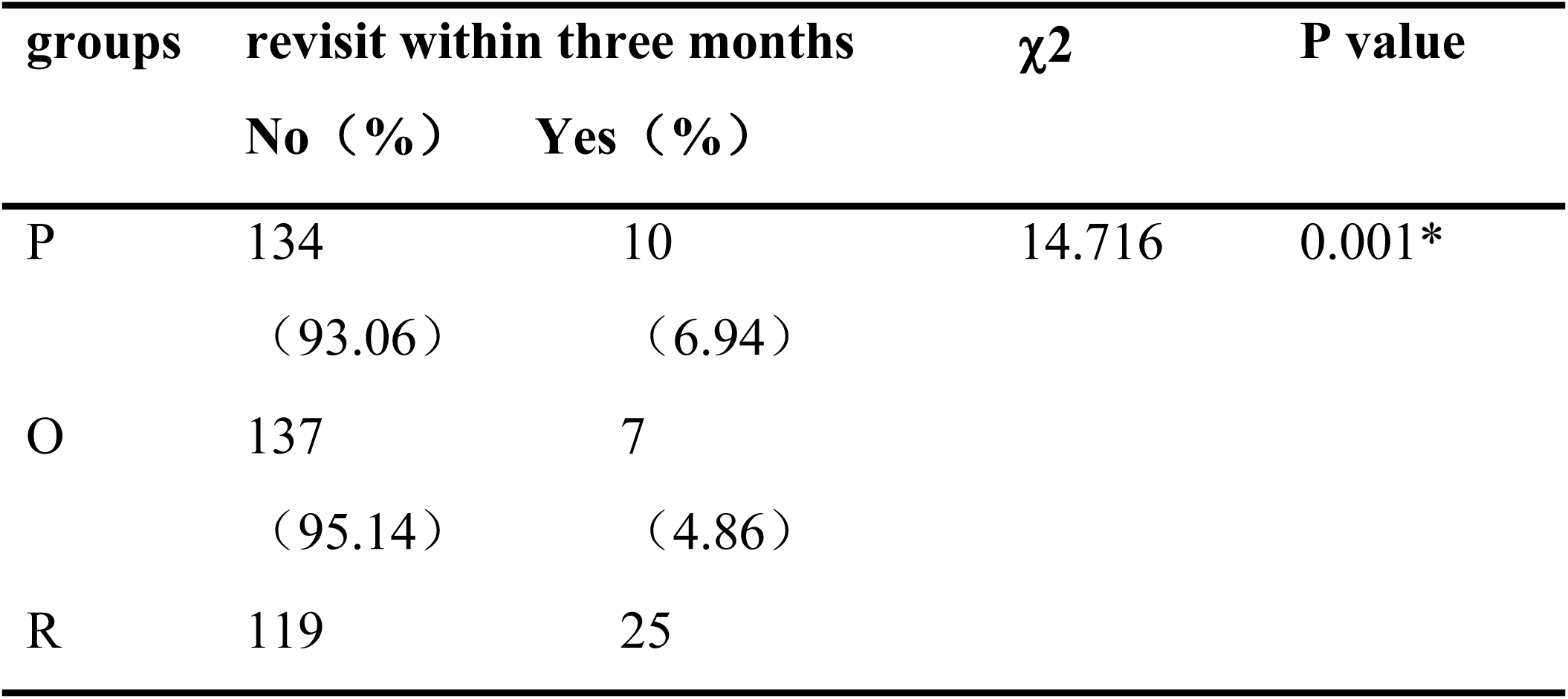
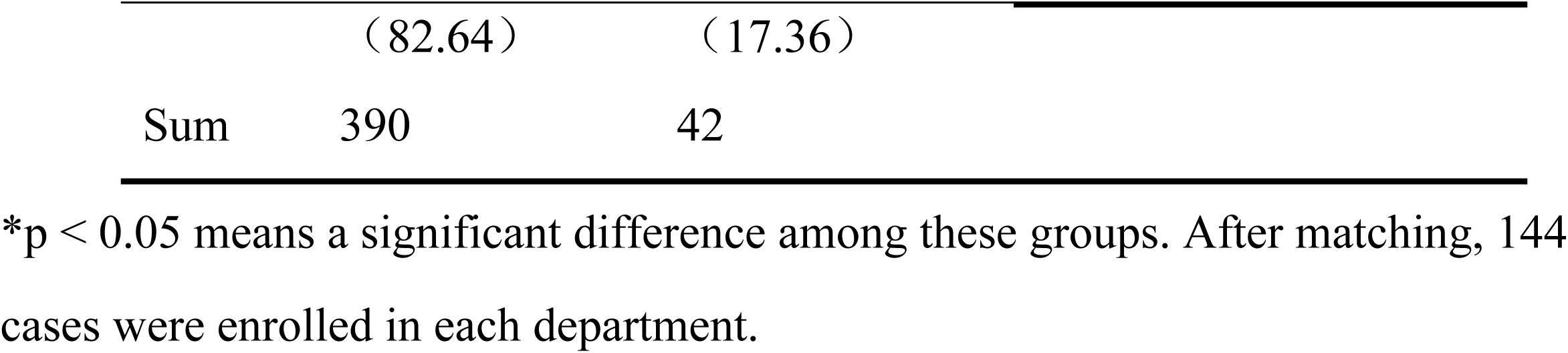
The rate of three months revisit after discharge in the three departments.

| groups | revisit within three months | | $\chi^2$ | P value |
| --- | --- | --- | --- | --- |
|  | No (%) | Yes (%) |  |  |
| P | 134<br>(93.06) | 10<br>(6.94) | 14.716 | 0.001* |
| O | 137<br>(95.14) | 7<br>(4.86) |  |  |
| R | 119 | 25 |  |  |

|  |  |  |
| --- | --- | --- |
|  | (82.64) | (17.36) |
| Sum | 390 | 42 |

\*p < 0.05 means a significant difference among these groups. After matching, 144 cases were enrolled in each department.

### The ODI

After PSM, there was no statistical difference in ODI at admission among the three groups (Table 6, P< 0.05). The ODI of the difference between the discharge and follow-up was statistically significant (Table 6, P< 0.05). The trend of ODI in the three groups is shown in Figure 2. Pair comparison showed that there was no statistically significant difference in ODI between group O and group P at the discharge and follow-up (P> 0.05), while there was statistically significant difference in ODI among group R, group P and group O at the discharge and follow-up (Table 7, P< 0.05).

**Figure 2.**
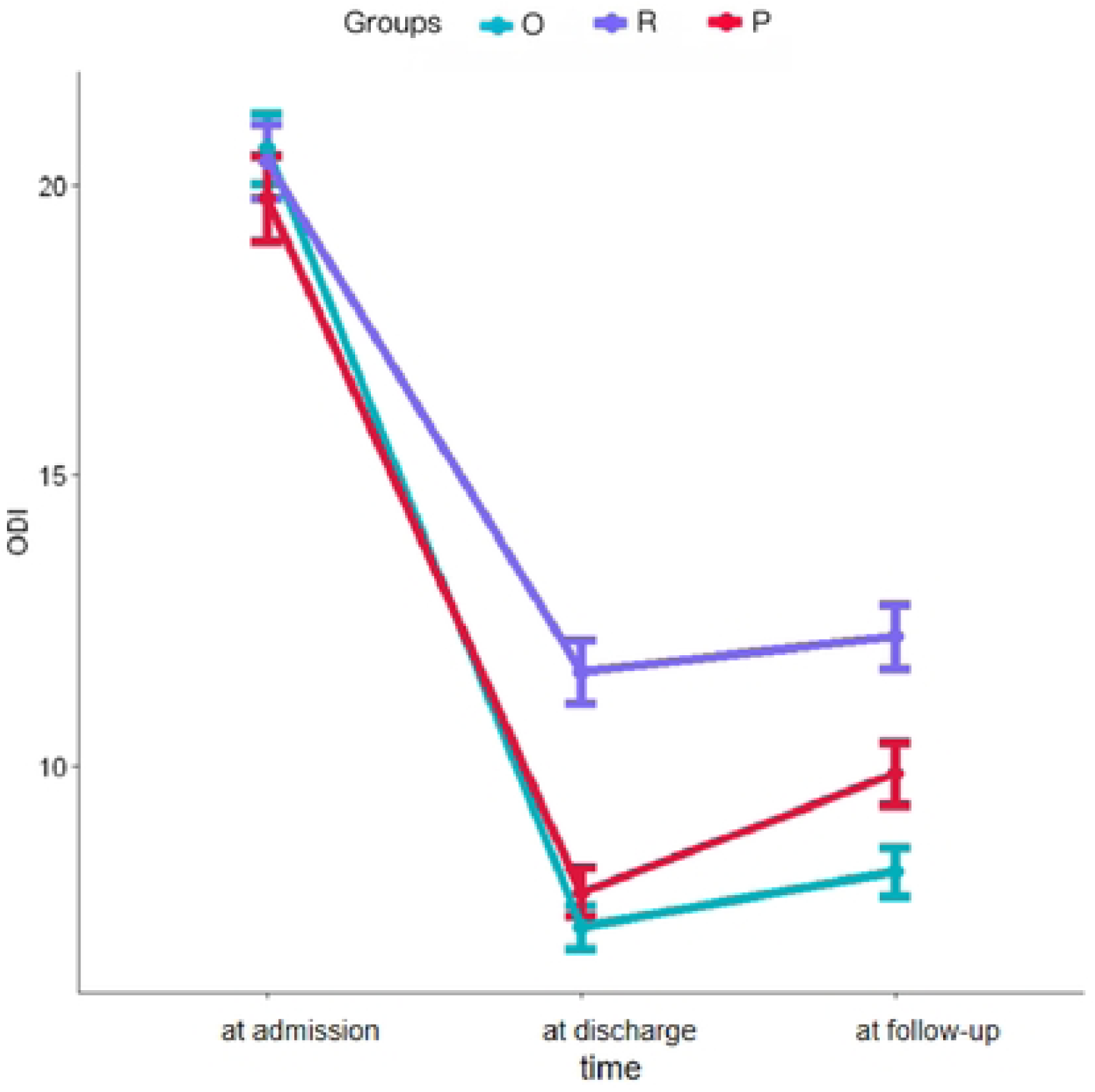
ODI trend plots in the three groups. Pair comparison showed that there was no statistically significant difference in ODI between group O and group P at the discharge and follow-up (P> 0.05).

**Table 6.** ODI of the three groups after matching.

| Characteristics | groups | P50 (P25-P75) | P value |
| --- | --- | --- | --- |
| ODI at admission(scores) | P | 19.50 (12.00-25.00) | 0.451 |
|  | O | 19.00 (15.00-25.00) |  |
|  | R | 19.00 (14.00-25.00) |  |
| ODI at discharge(scores) | P | 6.35 (3.80-10.43) | 0.000* |
|  | O | 6.00 (4.25-9.00) |  |
|  | R | 10.00 (7.00-14.00) |  |
| ODI at follow-up(scores) | P | 10.00 (4.00-14.00) | 0.000* |
|  | O | 7.00 (5.00-10.00) |  |
|  | R | 11.00 (8.00-15.00) |  |

\*p < 0.05 means a significant difference among these groups. After matching, 144 cases were enrolled in each department.

**Table 7.** Pairwise comparison of ODI in the three groups after matching.

| Characteristics | groups | groups | P value |
| --- | --- | --- | --- |
| ODI at | O | P | 1.000 |
| discharge(scores) | O | R | 0.000* |
|  | P | R | 0.000* |
| ODI at follow- | O | P | 0.053 |
| up(scores) | O | R | 0.000* |
|  | P | R | 0.000* |

\* $p < 0.05$ means a significant difference among these groups. After matching, 144 cases were enrolled in each department.

### The length of hospital stays and hospitalization cost

There was statistical difference in the length of hospital stay among the three departments (Table 8, P< 0.05), pair comparison showed that there was no significant difference in the length of hospital stay between group O and P (Table 9, P > 0.05), while there was significant difference between group R, group O and group P (Table 9, P < 0.05). There was no significant difference in length of stay between group O and P patients, and patients in group R had a longer hospital stay than those in group O and P.

**Table 8.**
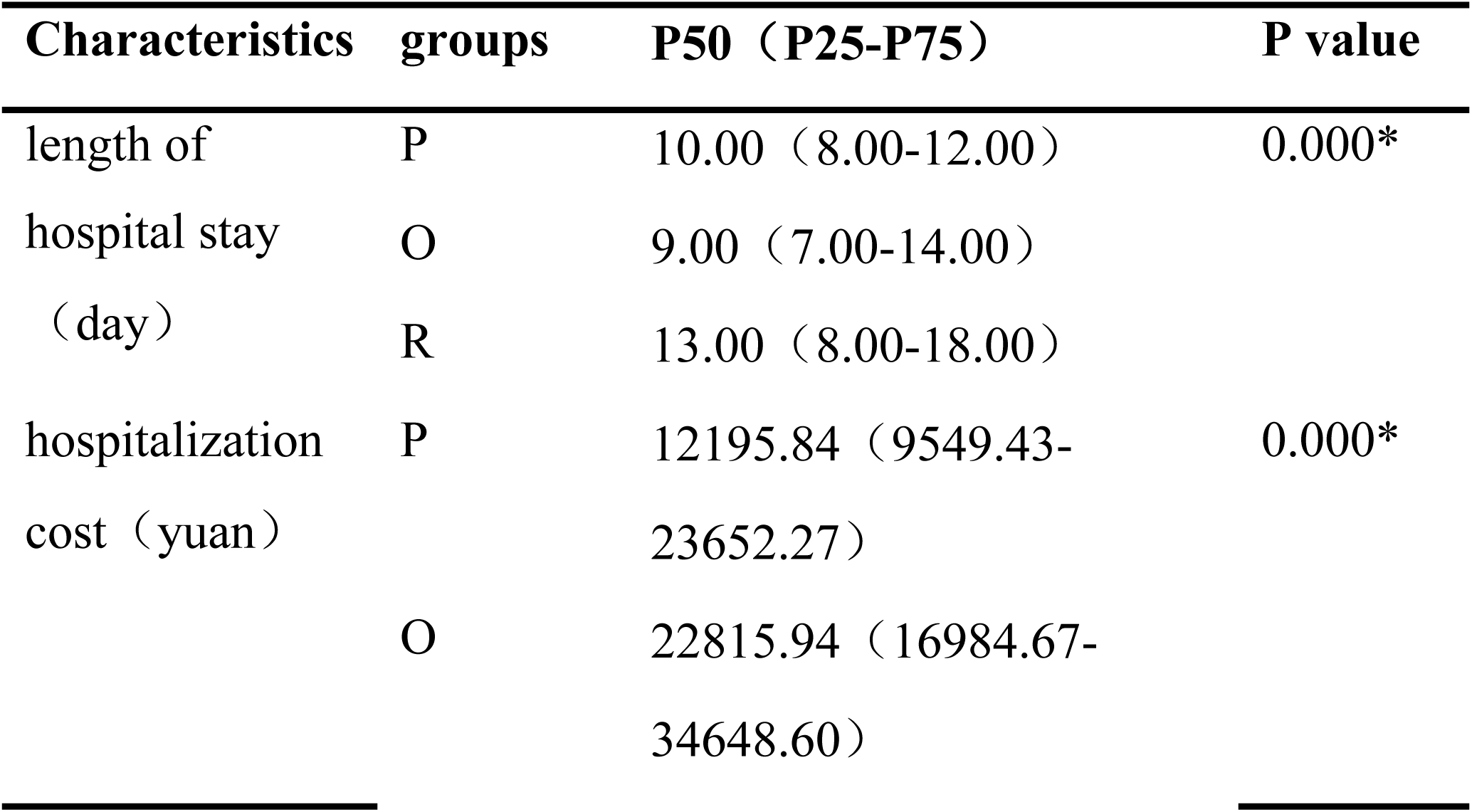
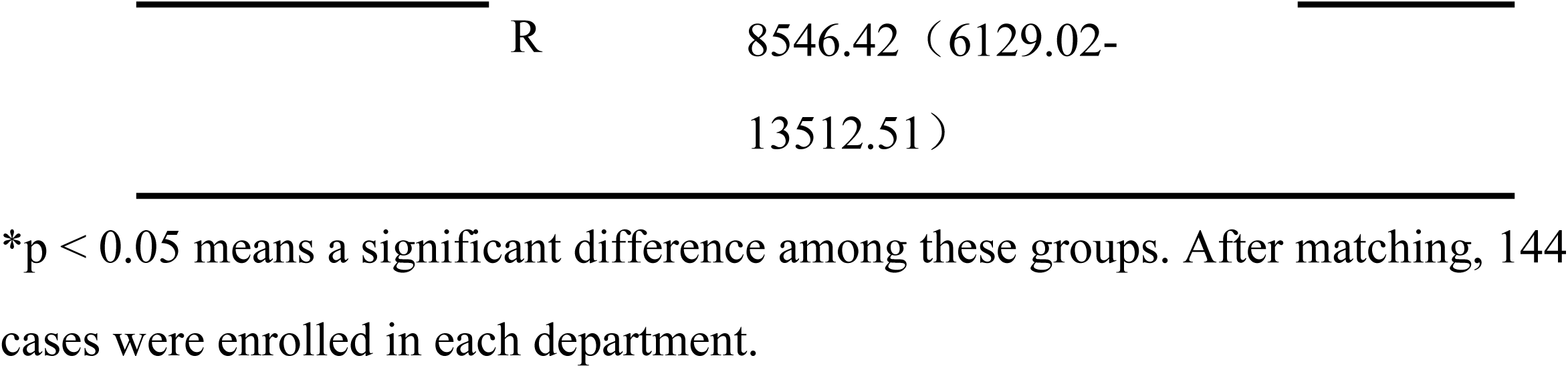
The length of hospital stays and the hospitalization cost in the three groups.

| Characteristics | groups | P50 (P25-P75) | P value |
| --- | --- | --- | --- |
| length of | P | 10.00 (8.00-12.00) | 0.000* |
| hospital stay | O | 9.00 (7.00-14.00) |  |
| (day) | R | 13.00 (8.00-18.00) |  |
| hospitalization | P | 12195.84 (9549.43- | 0.000* |
| cost (yuan) |  | 23652.27) |  |
|  | O | 22815.94 (16984.67- |  |
|  |  | 34648.60) |  |

|  |  |  |
| --- | --- | --- |
|  | R | 8546.42 (6129.02-13512.51) |

\*p < 0.05 means a significant difference among these groups. After matching, 144 cases were enrolled in each department.

**Table 9.** Pairwise comparison of the length of hospital stay among the three groups.

| Characteristics | groups | groups | P value |
| --- | --- | --- | --- |
| length of | O | P | 1.000 |
| hospital stay | O | R | 0.000* |
| (day) | P | R | 0.000* |

\*p < 0.05 means a significant difference among these groups.

There was statistical difference in the hospitalization cost among the three groups (Table 8, P< 0.05), pairwise comparison showed statistically significant difference in the hospitalization cost from the three departments (Table 10, P< 0.05), ranked as O>P>R.

**Table 10.**
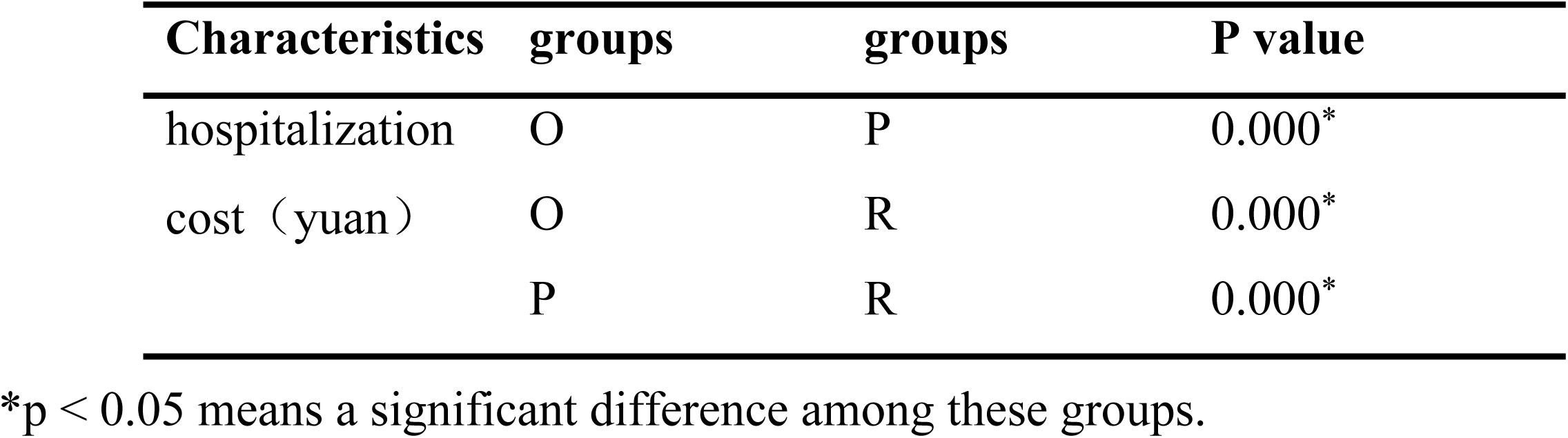
Pairwise comparison of hospitalization cost among the three groups.

| Characteristics | groups | groups | P value |
| --- | --- | --- | --- |
| hospitalization | O | P | 0.000* |
| cost (yuan) | O | R | 0.000* |
|  | P | R | 0.000* |

\*p < 0.05 means a significant difference among these groups.

## Discussion

The clinical treatment of LDH is diverse, and generally follows the stepped treatment of conservative treatment, minimally invasive procedures and surgical intervention[7]. Rehabilitation department, pain management department and orthopedic departments play corresponding roles within this approach. However, there has been no prior research comparing the efficacy of these three departments in LDH treatment.

Open surgery and minimally invasive surgery for LDH by orthopedics have shown definite efficacy in previous studies. As reported by Li Q et al.[16], the excellent rates of 93.5% for traditional laminotomy discectomy and 92.6% for endoscopic discectomy in a 2-year follow-up in 1100 patients, which showed the accuracy and stability of the efficacy of the two surgical methods. The meta-analysis of Feng F et al.[11] included 7 minimally invasive treatments in 29 studies: percutaneous endoscopic discectomy (PELD), standard open discectomy (SOD), standard open microsurgical discectomy (SOMD), chemical nucleolysis (CN), micro endoscopic discectomy (MED), percutaneous laser disc decompression (PLDD), automated percutaneous lumber discectomy (APLD). Through RCT study found the ranking of surgical success rate in LDH treatment as follows: PELD> SOD> SOMD> CN> MED> PLDD> APLD, of which PELD is one of the minimally invasive surgery commonly used by orthopedic surgeons. The effectiveness and long-term stability of orthopedic surgery treatment for LDH consistent with our study’s findings, where patients treated in orthopedic department had the highest discharge relief rate among the three groups.

In this study, our pain management department used five minimally invasive treatments for LDH, including Epidural Injection (EI), Percutaneous endoscopic lumber discectomy (PELD), Coblation Nucleoplasty (CA), Never Block (NB), and Radiofrequency Thermocoagulation (RF). EI is widely used among LDH patients, accounting for 62.62% in our study. According to the review of Smith CC et al. [17], EI for radicular pain in LDH can achieve 64% effective rate from follow-up to one year. Kennedy DJ et al. [18] followed up LDH patients after EI treatment for 5 years and showed that the optimal efficacy was observed at six months. Overall, various minimally invasive interventions in pain management for LDH show precise efficacy. However, compared to the excellent rates of patients treated by orthopedic surgery reported by Li Q et al.[16], the discharge remission rate of various pain management treatments is slightly lower. This consistent with our study’s results, where the discharge remission rate in the pain management group is lower than that in the orthopedic group. Demirel A et al.[19] proved that non-invasive spinal decompression therapy can improve both clinical symptoms and imaging results of patients with LDH. Ford J et al.[20] proposed personalized physical therapy for patients with low back pain, selecting various methods tailored to each patient’s specific condition, including patient education, manual therapy, massage, yoga, medicine and so on.

They found that with personalized physical therapy, 92.3% of patients completed the formulated intervention treatment, and using this personalized approach, significant pain relief was achieved within 5-8 weeks, compared to the 12 months typically required, which highlights the effectiveness of physical rehabilitation therapy.

However, there were still 6.7% of patients failed to complete their treatment plans. Thus, while the various physical therapy methods offered by rehabilitation departments can improve the symptoms of patients, they may not achieve curative effect as quickly as those seen in orthopedics or pain management department, as they do not direct removal of the protruding nucleus pulposus tissue or use medication.

Nguyen C A et al.[21] indicated in a prospective, randomized trial that a single injection of prednisolone can reduce low back pain for one month but does not improve low back pain after 12 months. Pennington Z et al.[22] compared the efficacy of EI and conservative treatment (drug and physical therapy) in disc-derived radiculopathy, found similar improvements in quality of life at three months for both treatments, but neither significantly improved the quality of life at six months. Based on the above researches, the treatment methods of O, P and R groups have instances of disease recurrence. Considering the principles of treatment methods in each department, consistent with our study results, the three-month revisits rate is ranked as R > P > O. As the treatment methods in the rehabilitation department do not involve the removal of the protruding nucleus pulposus or chemical/ drug therapys, the long-term stability of the efficacy is poor, so that the ODI index is easy to rebound after discharge.

Although there are no clear stipulation on the admission standards of LDH in the three departments, from our initial data analysis, we observed that the ODI index of patients in the three departments was O> P> R upon admission (Table 1), and the age of the three departments followed the order of P>O>R (Table 1), indicating that there are some default indicators for admission in each department. In our findings, orthopedics department achieved rapid symptom relief through open surgery or minimally invasive discectomy (70% at median discharge relief rate), pain management department achieved clinical symptom relief through various minimally invasive treatment methods (60% at median discharge relief rate), while rehabilitation department achieved certain clinical symptom resolution through various non-invasive treatment methods (50% at median discharge relief rate). Combined with the follow-up visit frequency within three months (R>P>O), we believe that each department is suitable for the treatment of different clinical stages of LDH.

In recent years, Multi-Disciplinary Treatment (MDT) has been increasingly widely used in clinical practice. Kintiraki E and Goulis DG[23] reported that multi- disciplinary management of gestational diabetes could provide more comprehensive blood sugar management during pregnancy. We believe that multi-disciplinary management constitutes an effective therapeutic approach of LDH. In previous studies, although the MDT pattern was not clearly proposed, the postoperative functional exercise has been valued by scholars. Zhang R et al.[24] reported that early functional exercise of passive and autonomic activities after minimally invasive surgery in LDH patients made the total effective rate after one year significantly higher than the control group (82.6% vs 71.7%) and improved the postoperative quality of life of LDH patients. A recent RCT by Demir S et al.[25] showed patients following spinal surgery with dynamic lumbar stabilization exercises are benefits in reducing pain, and ensuring faster return to work periods. Early rehabilitation exercise is beneficial for postoperative patient with LDH, whether surgical treatment or minimally invasive treatment.

In treatment method of the rehabilitation department, spine stable exercise (Stabilization Exercises) can effectively relieve pain and prevent fatigue in herniated disk patients[26]. Non-surgical Spinal Decompression Therapy improved both clinical symptoms and imaging results in patients with LDH[19]. Spinal Manipulation can significantly improve the range of motion in patients with non-acute lumbar radiculopathy, and significantly improve the ODI index[27]. Core Stability Exercise is more effective in reducing pain and increasing back-specific function in chronic back pain patients than General Exercise[28]. The application of these treatments in chronic pain in LDH has definite efficacy and definite improvement in the functional status of patients. Therefore, the rehabilitation department should be an important member of the MDT model of LDH patients. For patients in the acute phase, rehabilitation treatment cannot be performed due to severe pain. In this phase, significant pain relief can be achieved by surgical treatment of orthopedics or minimally invasive treatment of pain management department, further combined with the treatment of the rehabilitation department can promote the rapid recovery of the patients, and further improve the patient’s motor function[24, 29–31]. Therefore, orthopedics department and pain management departments should be considered as the constituent members of the MDT mode of LDH.

Regarding the treatment strategy of LDH, Wu PH et al.[7] proposed its general process by reviewing many relevant literatures. Patients diagnosed early with LDH often undergo conservative treatment or movement protection, typically managed by rehabilitation department. If these methods prove ineffective, pain management department may consider minimally invasive surgical options, especially for those with discogenic back pain. Patients with predominant disc collapse or gross instability may require orthopedic intervention such as disc replacement or vertebral fusion.

Combining the treatment process outlined by Wu PH et al. with our research findings, we propose a stepped treatment plan for patients with LDH, utilizing MDT therapy, to provide clinical treatment reference (Figure 3).

**Figure 3.**
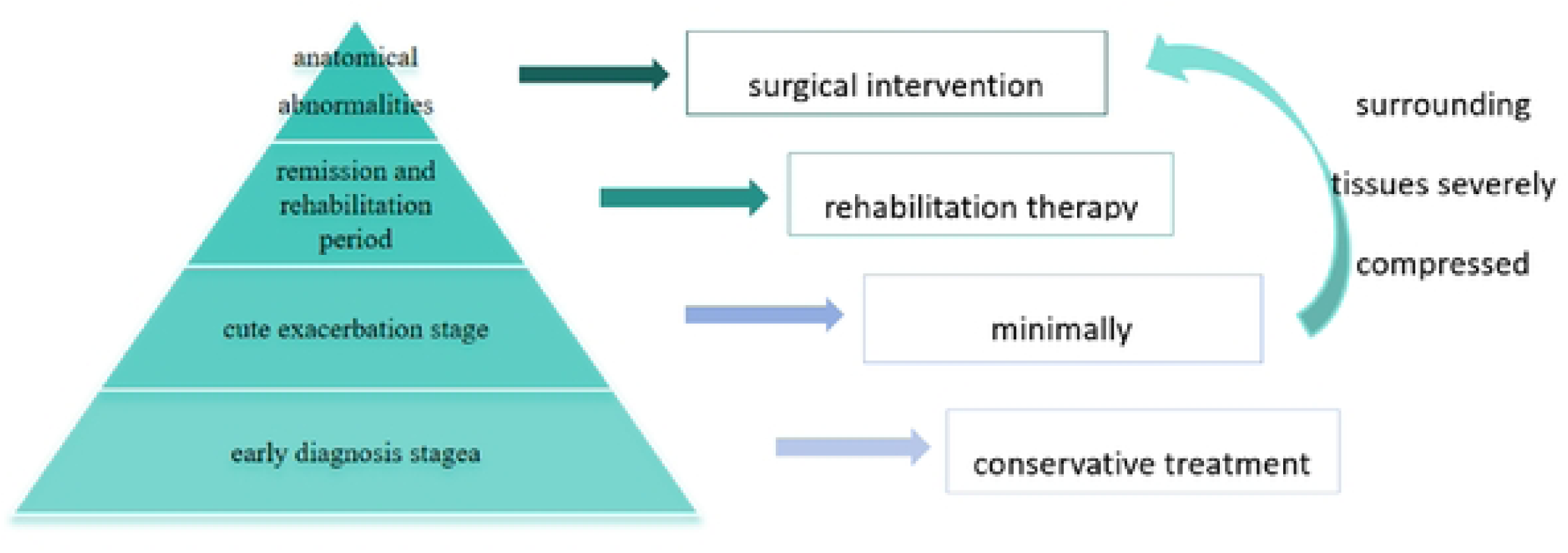
The stepwise treatments and MDT scheme for patients with LDH

In this scheme, the first step is for patients in the early diagnosis stage, where we recommend conservative treatment and activity protection, including bed rest, traction, exercise therapy, manual therapy, physical therapy, etc., with rehabilitation medicine as the focus. The second step is for patients in the acute exacerbation stage, we suggest medication therapy, primarily non-steroidal anti-inflammatory drugs (NSAIDs), with temporary use of opioid drugs if necessary. For patients with long-term or recurrent conditions, appropriate mood-improving medications may be used as well. Minimally invasive treatments such as EI, CA, NB, RF, PELD and related endoscopic techniques should be selected according to the patient’s condition. For patients in the acute exacerbation stage with severe compression of the surrounding tissues due to nucleus pulposus, direct entry into the fourth step involves intervertebral disc replacement surgery or discectomy and fusion surgery. The third step is for patients in remission and rehabilitation period, in this step, rehabilitation therapy and activity protection are recommended to avoid acute exacerbations and reduce patient re-visits. The fourth step is for patients with significant disc protrusion or anatomical abnormalities caused by vertebral instability. In this step, disc replacement surgery or disc excision followed by vertebral fusion surgery is recommended. In the stepped treatment, comprehensive rehabilitation department, pain management department, orthopedics department for comprehensive treatment. As with any study, there are some limitations to our study which need to be acknowledged. Firstly, we did not grade the severity of patient’s condition to compared the efficacy. Therefore, the evaluation of the efficacy may be biased.

Additionally, there is no record and statistical analysis of the complications of each treatment were performed in this study and further research is needed to get a more optimized path of LDH treatment. Despite these limitations, we believe that this study is clinically important significant since it compared the efficacy of LDH in the three departments, and used PSM method to match the baseline data in each group, which is different from the previous studies.

## Conclusions

The findings of this study suggest that in the treatment of LDH, the orthopedics, pain and rehabilitation departments could all achieve the relief of clinical symptoms. Through invasive or minimally invasive methods, orthopedics department was definite and stable efficacy, low short-term revisit rate. Through minimally invasive methods, pain management department was next to orthopedics department, while through non-invasive methods, rehabilitation department is slightly worse than above two. The treatment of LDH should follow the mode of ladder therapy and MDT. In the future, further studies should be erected to make the LDH patients get faster remission, better postoperative rehabilitation, to reduce the recurrence and re-visit rate.

## Data Availability

The data underlying the results presented in the study are available from the corresponding author Ling Ye.

## References

[1] H.Y. Nie, Y.B. Qi, N. Li, S.L. Wang, Y.X. Cao, Comprehensive comparison of therapeutic efficacy of radiofrequency target disc decompression and nucleoplasty for lumbar disc herniation: a five year follow-up, International orthopaedics 42(4) (2018) 843–849.

[2] D.J. Lawrence, W. Meeker, R. Branson, G. Bronfort, J.R. Cates, M. Haas, M. Haneline, M. Micozzi, W. Updyke, R. Mootz, J.J. Triano, C. Hawk, Chiropractic management of low back pain and low back-related leg complaints: a literature synthesis, Journal of manipulative and physiological therapeutics 31(9) (2008) 659–74.

[3] B. Zhang, H. Xu, J. Wang, B. Liu, G. Sun, A narrative review of non-operative treatment, especially traditional Chinese medicine therapy, for lumbar intervertebral disc herniation, Bioscience trends 11(4) (2017) 406–417.

[4] L. Zhao, L. Manchikanti, A.D. Kaye, A. Abd-Elsayed, Treatment of Discogenic Low Back Pain: Current Treatment Strategies and Future Options-a Literature Review, Current pain and headache reports 23(11) (2019) 86.

[5] H. Kanno, T. Aizawa, K. Hahimoto, E. Itoi, Minimally invasive discectomy for lumbar disc herniation: current concepts, surgical techniques, and outcomes, International orthopaedics 43(4) (2019) 917–922.

[6] J.N. Weinstein, T.D. Tosteson, J.D. Lurie, A.N. Tosteson, B. Hanscom, J.S. Skinner, W.A. Abdu, A.S. Hilibrand, S.D. Boden, R.A. Deyo, Surgical vs nonoperative treatment for lumbar disk herniation: the Spine Patient Outcomes Research Trial (SPORT): a randomized trial, Jama 296(20) (2006) 2441–50.

[7] P.H. Wu, H.S. Kim, I.T. Jang, Intervertebral Disc Diseases PART 2: A Review of the Current Diagnostic and Treatment Strategies for Intervertebral Disc Disease, International journal of molecular sciences 21(6) (2020).

[8] J. MacVicar, W. King, M.H. Landers, N. Bogduk, The effectiveness of lumbar transforaminal injection of steroids: a comprehensive review with systematic analysis of the published data, Pain medicine (Malden, Mass.) 14(1) (2013) 14–28.

[9] X. Yang, H. Wu, Analysis of the therapeutic effect and postoperative complications associated with 3-dimensional computed tomography navigation-assisted intervertebral foraminoscopic surgery in lumbar disc herniation in the elderly: a retrospective cohort study, Quantitative imaging in medicine and surgery 13(10) (2023) 7180–7193.

[10] Y.B. Liu, Y. Wang, Z.Q. Chen, J. Li, W. Chen, C.F. Wang, Q. Fu, Volume Navigation with Fusion of Real-Time Ultrasound and CT Images to Guide Posterolateral Transforaminal Puncture in Percutaneous Endoscopic Lumbar Discectomy, Pain physician 21(3) (2018) E265–e278.

[11] F. Feng, Q. Xu, F. Yan, Y. Xie, Z. Deng, C. Hu, X. Zhu, L. Cai, Comparison of 7 Surgical Interventions for Lumbar Disc Herniation: A Network Meta-analysis, Pain physician 20(6) (2017) E863–e871.

[12] Y. Yao, H. Zhang, J. Wu, H. Liu, Z. Zhang, Y. Tang, Y. Zhou, Comparison of Three Minimally Invasive Spine Surgery Methods for Revision Surgery for Recurrent Herniation After Percutaneous Endoscopic Lumbar Discectomy, World neurosurgery 100 (2017) 641–647.e1.

[13] Y. Yang, X. Yan, W. Li, W. Sun, K. Wang, Long-Term Clinical Outcomes and Pain Assessment after Posterior Lumbar Interbody Fusion for Recurrent Lumbar Disc Herniation, Orthopaedic surgery 12(3) (2020) 907–916.

[14] A. Bhatia, D. Flamer, P.S. Shah, S.P. Cohen, Transforaminal Epidural Steroid Injections for Treating Lumbosacral Radicular Pain from Herniated Intervertebral Discs: A Systematic Review and Meta-Analysis, Anesthesia and analgesia 122(3) (2016) 857–870.

[15] R. Smeets, A. Köke, C.W. Lin, M. Ferreira, C. Demoulin, Measures of function in low back pain/disorders: Low Back Pain Rating Scale (LBPRS), Oswestry Disability Index (ODI), Progressive Isoinertial Lifting Evaluation (PILE), Quebec Back Pain Disability Scale (QBPDS), and Roland-Morris Disability Questionnaire (RDQ), Arthritis care & research 63 Suppl 11 (2011) S158–73.

[16] Q. Li, Y. Zhou, Comparison of conventional fenestration discectomy with Transforaminal endoscopic lumbar discectomy for treating lumbar disc herniation:minimum 2-year long-term follow-up in 1100 patients, BMC musculoskeletal disorders 21(1) (2020) 628.

[17] C.C. Smith, Z.L. McCormick, R. Mattie, J. MacVicar, B. Duszynski, M.P. Stojanovic, The Effectiveness of Lumbar Transforaminal Injection of Steroid for the Treatment of Radicular Pain: A Comprehensive Review of the Published Data, Pain medicine (Malden, Mass.) 21(3) (2020) 472–487.

[18] D.J. Kennedy, P.Z. Zheng, M. Smuck, Z.L. McCormick, L. Huynh, B.J. Schneider, A minimum of 5-year follow-up after lumbar transforaminal epidural steroid injections in patients with lumbar radicular pain due to intervertebral disc herniation, The spine journal : official journal of the North American Spine Society 18(1) (2018) 29–35.

[19] A. Demirel, M. Yorubulut, N. Ergun, Regression of lumbar disc herniation by physiotherapy. Does non-surgical spinal decompression therapy make a difference? Double-blind randomized controlled trial, Journal of back and musculoskeletal rehabilitation 30(5) (2017) 1015–1022.

[20] J. Ford, A. Hahne, L. Surkitt, A. Chan, M. Richards. (2019).The Evolving Case Supporting Individualised Physiotherapy for Low Back Pain.Journal of clinical medicine, 8(9).

[21] C. Nguyen, I. Boutron, G. Baron, K. Sanchez, C. Palazzo, R. Benchimol, G. Paris, É. James-Belin, M.M. Lefèvre-Colau, J. Beaudreuil, J.D. Laredo, A. Béra-Louville, A. Cotten, J.L. Drapé, A. Feydy, P. Ravaud, F. Rannou, S. Poiraudeau. (2017).Intradiscal Glucocorticoid Injection for Patients With Chronic Low Back Pain Associated With Active Discopathy: A Randomized Trial. Annals of internal medicine, 166(8) ,547–556.

[22] Z. Pennington, M.A. Swanson, D. Lubelski, V. Mehta, M.D. Alvin, H. Fuhrman, E.C. Benzel, T.E. Mroz.(2020).Comparing the short-term cost-effectiveness of epidural steroid injections and medical management alone for discogenic lumbar radiculopathy. Clinical neurology and neurosurgery, 191, 105675.

[23] E. Kintiraki, D.G. Goulis, (2018),Gestational diabetes mellitus: Multi-disciplinary treatment approaches. Metabolism: clinical and experimental, 86,91–101.

[24] R. Zhang, S.J. Zhang, X.J. Wang. (2018).Postoperative functional exercise for patients who underwent percutaneous transforaminal endoscopic discectomy for lumbar disc herniation. European review for medical and pharmacological sciences, 22(1 Suppl) ,15-22.

[25] S. Demir, D. Dulgeroglu, A. Cakci. (2014). Effects of dynamic lumbar stabilization exercises following lumbar microdiscectomy on pain, mobility and return to work. Randomized controlled trial. European journal of physical and rehabilitation medicine, 50(6), 627–40.

[26] L.A.V. Ramos, B. Callegari, F.J.R. França, M.O. Magalhães, T.N. Burke, E.S.A. Carvalho, G.P.L. Almeida, J. Comachio, A.P. Marques.(2018). Comparison Between Transcutaneous Electrical Nerve Stimulation and Stabilization Exercises in Fatigue and Transversus Abdominis Activation in Patients With Lumbar Disk Herniation: A Randomized Study. Journal of manipulative and physiological therapeutics, 41(4), 323–331.

[27] S.H. Ghasabmahaleh, Z. Rezasoltani, A. Dadarkhah, S. Hamidipanah, R.K. Mofrad, S. Najafi. (2021).Spinal Manipulation for Subacute and Chronic Lumbar Radiculopathy: A Randomized Controlled Trial. The American journal of medicine, 134(1), 135–141.

[28] B.J. Coulombe, K.E. Games, E.R. Neil, L.E. Eberman. (2017).Core Stability Exercise Versus General Exercise for Chronic Low Back Pain. Journal of athletic training, 52(1), 71–72.

[29] A. Abdi, S.R. Bagheri, Z. Shekarbeigi, S. Usefvand, E. Alimohammadi. (2023). The effect of repeated flexion-based exercises versus extension-based exercises on the clinical outcomes of patients with lumbar disk herniation surgery: a randomized clinical trial.Neurological research, 45(1), 28–40.

[30] H. Chang, J. Xu, D. Yang, J. Sun, X. Gao, W. Ding,(2023).Comparison of full-endoscopic foraminoplasty and lumbar discectomy (FEFLD), unilateral biportal endoscopic (UBE) discectomy, and microdiscectomy (MD) for symptomatic lumbar disc herniation. European spine journal, 32(2), 542–554.

31. [31] W.C. Peul, W.B. van den Hout, R. Brand, R.T. Thomeer, B.W. Koes. (2008).Prolonged conservative care versus early surgery in patients with sciatica caused by lumbar disc herniation: two year results of a randomised controlled trial.BMJ (Clinical research ed.), 336(7657) ,1355–8.

